# PREDICTION OF ALL-CAUSE MORTALITY AND CARDIOVASCULAR EVENTS IN END STAGE RENAL DISEASE WITH CENTRAL HAEMODYNAMICS: A SYSTEMATIC REVIEW PROTOCOL

**DOI:** 10.1101/2022.07.30.22278242

**Authors:** Bárbara Letícia da Silva Guedes de Moura, Felipe Steinmacher Batista, Guilherme Luiz Rodrigues Ramajo, Gustavo Bueno Valente, João Vitor Scalon Estércio Rizzo, Tarik Radi Campos Maftoum, Izabel Galhardo Demarchi, Sérgio Seiji Yamada, Rogério Toshiro Passos Okawa, Jorge Juarez Vieira Teixeira

## Abstract

**Background:** Given the high rates of adverse events in End-stage renal disease (ESRD) patients and as the applications of central haemodynamics (central blood pressure, augmentation index, pulse wave velocity) in dialysis patients have not been fully clarified, we, therefore, will perform a systematic review of central haemodynamics as a predictor of total and cardiovascular mortality and morbidity in the ESRD population.

**Methods:** This protocol is reported according to the PRISMA-P guideline. The databases PubMed, EMBASE, LILACS, Web of Science, and Google Scholar system will be searched and double screening for prospective studies that assessed the prospective association of central haemodynamics parameters with at least one of the pre-specified outcomes in ESRD patients. Discrepancies will be resolved through consensus. A modified version of the Newcastle-Ottawa Quality Assessment Scale of cohort studies will be used. We intend to use the random-effects model, considering at least moderate heterogeneity between studies. If data allow, we will perform a sensitivity analysis to explore potential sources of between-study heterogeneity.

**Discussion:** This systematic review will summarize current evidence about central haemodynamics measures in the ESRD patients and clarify the incremental prognostic value of this diagnostic tool in these patients. Therefore, this systematic review will also be displayed important knowledge gaps in the current literature regarding the possible utility of central haemodynamics indices in ESRD patients.

**Systematic review registration:** This systematic review protocol was submitted to the PROSPERO international registry of systematic reviews (Registration number: CRD42022260824).

**Strengths and limitations of this study:**

→ The PROSPERO registry helps to promote and maintain transparency in the process and to assist in minimizing the risk of bias. Four databases and one additional grey literature source helps ensure more complete coverage of the topic. All the review phases will be peer-reviewed and will be validated by methodological and clinical experts.
→ Central (aortic, carotid) pressures are pathophysiologically more relevant than peripheral pressures for the pathogenesis of cardiovascular (CV) disease. There is a cumulative increase in CVD during the transition from CKD to ESRD: as patients initiate dialysis, there are significant changes in blood pressure, volume status, and circulating solutes, all of which may contribute to increased risk of CVD. Studies did not further analyze the ability of central pressures and indices to predict future events in separated stages of CKD, especially in End-stage renal disease patients (ESRD) patients.
→ This study has limitations. Because heterogeneity is quite common in these systematic reviews, there may be limited scope for meta-analysis. Possibly, the spectrum of patients in different research sites is the main factor, in most cases, but heterogeneity in associations between studies may be difficult to explain.
→ This systematic review will evaluate the evidence related to the Central Haemodynamics (central blood pressure, augmentation index, pulse wave velocity) incremental prognostic value in ESRD patients, which may lower morbidity and mortality and influence its use as a prognostic marker for ESRD patients.

## Background

End-stage renal disease (ESRD) is a rapidly increasing global health and health care burden. The number of people receiving renal replacement therapy (RRT) exceeds 2.5 million and is projected to double to 5.4 million by 2030 (1). Data from the US Renal Data System (USRDS) reported that among CKD patients who die, 91% have a diagnosis of cardiovascular disease (CVD) overall, 57% have a diagnosis of coronary (CAD), and 63% have a diagnosis of Heart Failure (HF).

Although classical Framingham risk factors remain important, there is increasing evidence to support renal-specific and dialytic disturbances increases the CV risk in this population, such as sodium/fluid balance, hyperuricemia, inflammation, vascular calcification, and abnormal bone mineral metabolism, and these complex interaction results in a high prevalence of abnormalities of cardiac structure and function in patients with ESRD (2,3).

Central (aortic, carotid) pressures are pathophysiologically more relevant than peripheral pressures for the pathogenesis of cardiovascular (CV) disease. Central haemodynamics (central/aortic blood pressure, augmentation index, pulse wave velocity) can now be reliably assessed non-invasively with a number of relatively inexpensive devices (4). While several studies have shown the ability of central pressures and indices to predict future events, findings have not always been consistent in the ESRD patients (5). Studies did not further analyze central haemodynamics in different stages of CKD, especially in ESRD patients. There is a cumulative increase in CVD during the transition from CKD to ESRD: as patients initiate dialysis, there are significant changes in blood pressure, volume status, and circulating solutes, all of which may contribute to increased risk of CVD (3,6,7).

Given the high rates of adverse events in dialysis patients and as the applications of central haemodynamics in dialysis patients have not been fully clarified, we, therefore, will perform a systematic review of central haemodynamics as a predictor of total and cardiovascular mortality and morbidity in the ESRD population.

## Methods/Design

This protocol is reported according to the Preferred Reporting Items for Systematic Reviews and Meta-Analyses Protocols (PRISMA-P) (8) and the corresponding checklist used (Supplementary file 1). This includes details of inclusion and exclusion criteria, a data extraction tool, and analytical methods. This systematic review protocol was submitted to the PROSPERO international registry of systematic reviews (Registration number: CRD42022260824).

### Review question

Is the Central Haemodynamics (central blood pressure, augmentation index, pulse wave velocity) a better prognostic marker than peripheral measures (brachial blood pressure/ no control) in the End Stage Renal Disease?

P: End-stage renal disease patients

E: Central Haemodynamics (Central blood pressure, Augmentation Index and/or Pulse Wave Velocity)

C: Peripheral (brachial) blood pressure / no restriction O: All-cause mortality, cardiovascular events

S: Prospective studies

### Searches

MEDLINE (using the PubMed interface), Web of Science, LILACS (Latin American and Caribbean Literature in Health Sciences) and Google Scholar (grey literature) databases and reference lists of eligible studies will be searched for publications until the search date. The Search strategy were determined independently by three authors (BLSG, RTPO and JJVT). The search will be limited to original research reports in peer-reviewed journals with no language or publication date restriction.

### Search strategy

The search strategy will be developed first for the MEDLINE database and the effectiveness will be tested and refined accordingly, using the keywords ‘Central Aortic Blood Pressure, ‘Pulse Wave Analysis’, ‘Vascular stiffness’, ‘chronic kidney disease’, ‘hemodialysis’. A detailed draft for the search strategy to be used is detailed in Supplementary file 2.

### Eligibility criteria

All longitudinal studies that assessed the prospective association of one or more index of central haemodynamics (central blood pressure - cBP, Augmentation Index - Aix, or Pulse Wave Velocity - PWV) with: (a) at least one of the pre-specified outcomes in ESRD (>18 years) population and (b) in accordance with ethical standards, will be included and (c) Patients data with CKD in different stages, (not only ESRD or which one that not reported the results separately according to the CKD stage) and (d) Abstracts, reviews, conference proceedings or letters to the editor will be excluded.

Search results from each database will be combined and duplicates will be removed before the screening. References will be aggregated into a reference manager tool (EndNote Web) (9). Additional papers will be identified by searching the reference lists of relevant articles and their citation metrics.

A draft for the Eligibility and Data Extraction Form to be used is detailed in Supplementary file 3.

### Outcomes and Measures of effect

The primary outcome will be all-cause mortality whereas secondary outcomes will be a composite cardiovascular (endpoint includes: CV mortality, Coronary Heart Disease (CHD) events (Myocardial infarction (MI), unstable angina, angina/ischemia requiring emergent hospitalization or revascularization), Heart Failure (HF) hospitalization, new onset Atrial Fibrillation (AF), life threatening arrhythmia, recorded automatic implantable cardioverter defibrillator (AICD) shocks, stroke, transit ischemic attack, peripheral arterial disease (PAD) with arterial revascularization procedure). Tertiary outcomes will be any individual secondary endpoint not included in the composite cardiovascular.

We will provide a narrative synthesis of the findings from the included studies. We will provide summaries of intervention effects for each study by calculating risk ratios (for dichotomous outcomes) or standardized mean differences (for continuous outcomes) from the data presented in the published studies or obtained from study authors.

### Study reviews and appraisals

Initial title and abstract screening will be performed using Rayyan software (10) and full texts of selected articles will be retrieved and double screened for eligibility and data extraction by six researchers working independently (BLSGM, FSB, GLRR, GBV, JVSER, TRCM). Discrepancies will be resolved through consensus or the opinion of a third review author (JJVT or RTPO) will be sought. For multiple study publications from the same patient cohort, we will choose the study with the largest number of cases, or which one that reported the results separately according to the CKD stage, once there are many studies whose results of patients with early-to-moderate CKD stage are reported together with ESRD patients, which makes it impossible to correctly analyze these subgroups. For studies that presented different outcomes, we will be extracted outcomes from both publications.

Reasons for study exclusion will be documented. Data extraction will be validated by methodological and clinical experts (IGD, JJVT and RTPO, SSY, respectively). Also, information reported in the grey literature will be sought. We will report our search terms as an appendix to published studies.

### Data extraction

We will extract the following data: (a) citation details (title, type, and year of publication); (b) study details (name, region, and design of the study, sample size); (c) participant details including demographics (age, sex, ethnicity, and follow-up duration), clinical (hypertension, diabetes mellitus-DM, dyslipidemia, smoking status, and known CVD), and laboratory variables; (d) exposure details (hardware and the software used for the measures acquisition and analysis); (e) details of outcomes, including how these were ascertained; (f) statistical methods used, as well as statistics related to the association of interest, and (g) information related to adjustment for potential confounders.

If samples are reported at multiple time points within a study, then each one will be considered individually. If these data are not available, we will extract the association measures that were used. The relevant authors will not be contacted for further information.

Quality assessment, data synthesis, and exploration of heterogeneity

### Risk of bias (quality) assessment

A modified version of the Newcastle-Ottawa Quality Assessment Scale of cohort studies (NOS) (11) will be used to assess the quality of included papers (Supplementary file 4). Studies will not be excluded from the main meta-analysis based on the risk of bias.

### Strategy for data synthesis

Statistical analyses will be performed using Review Manager 5.4 software. This information will be summarized graphically in the final review document. The degree of heterogeneity will be assessed using the Higgins Thompson I^2^ test (12) and Cochran’s Q test (13). If the included studies are sufficiently homogeneous, a quantitative synthesis using random-effect meta-analysis will be used to pool the results of the association of interest. For presenting the results, we will likely use the HR (95%CI) as an effect estimate and will present the data graphically in forest plots. If data allow, we will perform a sensitivity analysis to explore potential sources of between-study heterogeneity: (a) study quality, (b) variation in exposure measurement, and (c) variation in CV outcomes definition. Possible publication bias will be assessed using a Funnel plot (14). We will assess evidence of publication bias using Egger’s weighted regression method for continuous outcomes and Begg’s rank correlation test for dichotomous outcomes.

## Discussion

This systematic review will summarize current evidence about the incremental prognostic value of central haemodynamics measures in the ESRD patients. Studies reported associations between central haemodynamics measures and all-cause mortality and cardiovascular events, but, is not fully understood the potential prognostic value of this diagnostic tool in ESRD patients. Therefore, this systematic review will also be displayed important knowledge gaps in the current literature regarding the possible utility of central haemodynamics indices in ESRD patients.

Though hemodialysis (HD) should theoretically improve cardiovascular function by correcting fluid overload and small molecule accumulation, cardiovascular mortality continues to be disproportionately high in the HD population (1,2). The impact of renal replacement therapy (RRT) on cardiovascular function and injury is not well understood and may inadvertently be contributing to the accelerated development of type 4 cardiorenal syndrome (CRS): CKD leading to an impairment of cardiac function. The unique physiology of cardiovascular abnormalities in dialysis patients remains poorly understood and several more recently recognized factors, including altered lipid metabolism and accumulation of gut microbiota-derived uremic toxins, also affect cardiovascular function in the context of renal failure (15).

A crucial remaining question is whether central BP offers significant improvement in cardiovascular risk assessment and stratification compared with brachial (peripheral) BP. Several studies showed superiority of central compared with brachial BP; yet these findings have not always been consistent.

This systematic review will provide essential data regarding central haemodynamics as prognostic indicators of mortality and CV events in End-stage renal disease patients. The clinical implications of this work relate to the current status of diagnosis and treatment of heart disease in patients receiving dialysis.

## Availability of data and materials

The datasets generated and/or analyzed during the current study are available from the corresponding author on reasonable request.

## Supporting information

Supplementary_file_1_PRISMA-P_Checklist

Supplementary_file_2_Search_strategy

Supplementary_file_3_Eligibility_and_Data_Extraction_Forms

Supplementary_file_4_Quality_Assesment_Scale

## Data Availability

All data generated or analyzed during this study are included in this published article [and its supplementary information files].

## List of Abbreviations

Aix: Augmentation Index
AF: Atrial fibrillation
AICD: Automatic implantable cardioverter defibrillator
BP: blood pressure
CHD: Coronary heart disease
CKD: chronic kidney disease
CRS: Cardiorenal syndrome
CV: Cardiovascular
CVD: Cardiovascular disease
DM: Diabetes mellitus
ESRD: End-stage renal disease
HD: Hemodialysis
HF: Heart failure
LILACS: Latin American and Caribbean Literature in Health Sciences
MI: Myocardial infarction
NOS: Newcastle-Ottawa Quality Assessment Scale of cohort studies
PRISMA-P: Preferred Reporting Items for Systematic Reviews and Meta-Analyses Protocols
PROSPERO: International registry of systematic reviews
PP: Pulse pressure
PWV: Pulse wave velocity
RRT: Renal replacement therapy
UC: Uremic cardiomyopathy

## DECLARATIONS

### Ethics approval and consent to participate

Not applicable.

### Consent for publication

Not applicable.

### Authors’ contributions

BLSGM has prepared this manuscript with clinical and methodological support from RTPO and JJVT, respectively. This manuscript has been reviewed by FSB, GLRR, GBV, JVSER, TRCM, IGD, SSY, RTPO and JJVT and edited by IGD, SSY, RTPO and JJVT. BLSGM is the corresponding author and has registered the protocol with the PROSPERO database. All authors agreed to be accountable for all aspects of this work and approved the final submission.

## Supplementary information

Supplementary file 1: PRISMA-P Checklist.

Supplementary file 2: Draft Search strategy.

Supplementary file 3: Eligibility Form / Data Extraction Form.

Supplementary file 4: Modified NOS Risk of Bias assessment.

## REFERENCES

1. USRDS ADR. Epidemiology of kidney disease in the United States. National Institutes of Health, National Institute of Diabetes and Digestive and Kidney Diseases, Bethesda, MD. 2019 november. Available from: https://www.usrds.org/annual-data-report/

2. Bansal N. Evolution of Cardiovascular Disease During the Transition to End-Stage Renal Disease. Semin Nephrol. 2017 Mar;37(2):120–131. eng. doi:10.1016/j.semnephrol.2016.12.002. Cited in: Pubmed; PMID 28410646.

3. Ibrahim MK, Kamal OMM, Hassan MS, Khalifa MMM. Interdialytic Weight Gain and Its Relation to Outcome among Patients on Maintenance Hemodialysis. QJM: An International Journal of Medicine. 2020 2020-03-01;113(Supplement_1). doi:10.1093/qjmed/hcaa052.020.

4. Battistoni A, Michielon A, Marino G, Savoia C. Vascular Aging and Central Aortic Blood Pressure: From Pathophysiology to Treatment. High Blood Press Cardiovasc Prev. 2020 Aug;27(4):299–308. doi: 10.1007/s40292-020-00395-w. Epub 2020 Jun 22. PMID: 32572706.

5. Sabovic M, Safar ME, Blacher J. Is there any additional prognostic value of central blood pressure wave forms beyond peripheral blood pressure? Curr Pharm Des. 2009;15(3):254–66. doi: 10.2174/138161209787354249. PMID: 19149615.

6. Briet M, Boutouyrie P, Laurent S, London GM. Arterial stiffness and pulse pressure in CKD and ESRD. Kidney Int. 2012 Aug;82(4):388–400. doi: 10.1038/ki.2012.131. PMID: 22534962.

7. Protogerou AD, Papaioannou TG, Lekakis JP, Blacher J, Safar ME. The effect of antihypertensive drugs on central blood pressure beyond peripheral blood pressure. Part I: (Patho)-physiology, rationale and perspective on pulse pressure amplification. Curr Pharm Des. 2009;15(3):267–71. doi: 10.2174/138161209787354267. PMID: 19149617.

8. Moher D, Shamseer L, Clarke M, Ghersi D, Liberati A, Petticrew M, Shekelle P, Stewart LA. Preferred Reporting Items for Systematic Review and Meta-Analysis Protocols (PRISMA-P) 2015 statement. Syst Rev. 2015; 4(1):1. doi: 10.1186/2046-4053-4-1.

9. ENDNOTE WEB McKinney, A. EndNote Web: Web-Based Bibliographic Management. Journal of electronic resources in medical libraries, 2013 10(4), 185–192.

10. Ouzzani M, Hammady H, Fedorowicz Z, Elmagarmid A. Rayyan—a web and mobile app for systematic reviews. Systematic reviews, 2016. 5(1), 210.

11. Wells G, Shea B, O’Connell D, Peterson J, Welch V, Losos M, Tugwell P. Newcastle-Ottawa quality assessment scale cohort studies. 2014. 2015-11-19]. Available on: http://www.ohri.ca/programs/clinical_epidemiology/oxford.asp.

12. Higgins JP, Thompson SG, Deeks JJ, et al. Measuring inconsistency in meta-analyses. BMJ 2003;327:557–60. Higgins JP, Thompson SG. Quantifying heterogeneity in a metaanalysis. Stat Med 2002;21:1539–58.

13. Cochran WG. The comparison of percentages in matched samples. Biometrika, 1950. 37(3/4), 256–266.

14. Sterne, JA, Sutton AJ, Ioannidis JP, Terrin N, Jones DR, Lau J, Tetzlaff J. Recommendations for examining and interpreting funnel plot asymmetry in meta-analyses of randomised controlled trials. BMJ, 2011 343.

15. Ahmadmehrabi S, Wilson Tang WH. Hemodialysis-induced cardiovascular disease. Seminars in Dialysis. 2018; 31:258–267.

