## Supplementary_file_2_Search_strategy for "PREDICTION OF ALL-CAUSE MORTALITY AND CARDIOVASCULAR EVENTS IN END STAGE RENAL DISEASE WITH CENTRAL HAEMODYNAMICS: A SYSTEMATIC REVIEW PROTOCOL"

Draft search strategy to be used for the PubMed online database

| **Search query:** Is the Central Haemodynamics measures (central blood pressure, augmentation index, pulse wave velocity) a better prognostic marker than peripheral measures (brachial blood pressure/ no control) in the End Stage Renal Disease? | | | |
| --- | --- | --- | --- |
| **Sources searched: PubMed** | | | |
| **Limits: None**   - Date: 04/07/2021   Search number, Query, Sort By, Search details, Results | | | |
| Block 1 | 1 | (Vascular Stiffness[MeSH Terms]) | 5953 |
|  | 2 | (Pulse Wave Analysis[MeSH Terms]) | 4107 |
|  | 3 | (Central Aortic Blood Pressure[MeSH Terms]) | 792 |
|  | 4 | (Augmentation index [All Fields]) | 10109 |
|  | 5 | (Blood Pressure Monitoring, Ambulatory[MeSH Terms]) | 9822 |
| Block 2 | 6 | (Renal Dialysis[MeSH Terms]) | 113422 |
|  | 7 | (Kidney diseases[MeSH Terms]) | 511458 |
| Block 3 | 8 | (Risk Factors[MeSH Terms]) | 822527 |
|  | 9 | (Risk Assessment[MeSH Terms]) | 267639 |
|  | 10 | (Prognosis[MeSH Terms])) | 1625330 |
|  | 11 | (Death[MeSH Terms]) | 148153 |
|  | 12 | (Mortality[MeSH Terms]) | 380973 |
|  | 13 | OR (Morbidity[MeSH Terms]) | 555090 |
| 14  (1 OR 2 OR 3 OR 4 OR 5) | | ((((Vascular Stiffness[MeSH Terms]) OR (Pulse Wave Analysis[MeSH Terms])) OR (Central Aortic Blood Pressure[MeSH Terms])) OR (Augmentation index)) OR (Blood Pressure Monitoring, Ambulatory[MeSH Terms]) | 26587 |
| 15  (6 OR 7) | | (Renal Dialysis[MeSH Terms]) OR (Kidney diseases[MeSH Terms]) | 556759 |
| 16  (8 OR 9 OR 10 OR 11 OR 12 OR 13) | | (((((Risk Factors[MeSH Terms]) OR (Risk Assessment[MeSH Terms])) OR (Prognosis[MeSH Terms])) OR (Death[MeSH Terms])) OR (Mortality[MeSH Terms])) OR (Morbidity[MeSH Terms]) | 3050698 |
| **17**  **(14 AND 15 AND 16)** | | **((((((Vascular Stiffness[MeSH Terms]) OR (Pulse Wave Analysis[MeSH Terms])) OR (Central Aortic Blood Pressure[MeSH Terms])) OR (Augmentation index)) OR (Blood Pressure Monitoring, Ambulatory[MeSH Terms])) AND ((Renal Dialysis[MeSH Terms]) OR (Kidney diseases[MeSH Terms]))) AND ((((((Risk Factors[MeSH Terms]) OR (Risk Assessment[MeSH Terms])) OR (Prognosis[MeSH Terms])) OR (Death[MeSH Terms])) OR (Mortality[MeSH Terms])) OR (Morbidity[MeSH Terms]))** | **965** |
