## Supplementary_file_3_Eligibility_and_Data_Extraction_Forms for "PREDICTION OF ALL-CAUSE MORTALITY AND CARDIOVASCULAR EVENTS IN END STAGE RENAL DISEASE WITH CENTRAL HAEMODYNAMICS: A SYSTEMATIC REVIEW PROTOCOL"

| **1. Reference details** | |
| --- | --- |
| Ref ID |  |
| Reference citation |  |
| First author |  |
| Year of publication |  |
| Title of the paper |  |
| Type of publication | Paper ( ) Abstract ( ) Editorial ( )  Letter to Editor ( ) Review( ) other:___________________________________ |
| Assessor’s ID | 1 ( ) 2( ) 3( ) 4( ) 5( ) 6( ) |
| Date | __/__/__ |

| **2. Study Eligibility** | | | |
| --- | --- | --- | --- |
| Inclusion of the study | Yes( ) | No( ) | Unclear( ) |
| **Eligibility criteria** | **Reason for exclusion (if excluded)** | **YES** | **NO** |
| Type of publication: Paper | Reviews, abstracts, letters, conference proceeding. Other: __________________ | ( ) | ( ) |
| Population: End Stage Renal Disease | Unrepresentative sample of the population: __________________________________ | ( ) | ( ) |
| Exposure: Central Haemodynamics (Central blood pressure, Augmentation Index and/or Pulse wave velocity) | Ineligible exposure _____________________ | ( ) | ( ) |
| Outcomes: All cause mortality / CV events | Ineligible outcome (Different from outcomes considered in this review) ______ | ( ) | ( ) |
| Type of study: Prospective (Cohort) study | Ineligible study design (describe the type of study):_______ ____________ | ( ) | ( ) |
| **Other reason for exclusion:** | _____________________ | ( ) | ( ) |
|  | Duplicate | ( ) | ( ) |
| **Notes:** | | | |

**Additional information**

Notes:

**DO NOT PROCEED IF PAPER EXCLUDED FROM REVIEW**

**Data Extraction Form:** PREDICTION OF ALL-CAUSE MORTALITY AND CARDIOVASCULAR EVENTS IN END STAGE RENAL DISEASE WITH CENTRAL HAEMODYNAMICS: A SYSTEMATIC REVIEW PROTOCOL

| **1. Study details** | | |
| --- | --- | --- |
| Ref ID |  | |
| Study (cohort) name |  | |
| Study design | Prospective (cohort) study | |
| Region/country |  | |
| Sample size |  | |
| Mean (range) age | Mean ± SD: | Range (IQR): |
| Sex | Male, n (%): | Female, n (%): |
| Ethnicity |  |  |
| Follow-up duration (months) |  | |
| Clinical variables (n, %) |  | |
| Hypertension |  | |
| Diabetes mellitus |  | |
| Dyslipidemia |  | |
| Smoking status |  | |
| Known CVD |  | |
| Ethics: | Does the study include/mention a statement regarding the ethical approval, or adherence to an appropriate standard (such as the declaration of Helsinki)? YES( ) NO( ) | |

| **2. Exposure details** | | | |
| --- | --- | --- | --- |
| Central haemodynamics measure (Central Blood pressure, Aix, Pulse wave velocity): | Hardware: | Software: | Procedure: |
| Notes: | | | |

| **3. Outcomes’ details** | |
| --- | --- |
| Type of outcomes used in this review | Outcomes reported in this study (specify the outcome, n, %) |
| **Primary outcome(s)**  All-cause mortality |  |
| **Secondary outcome (s)**  Composite cardiovascular (CV) |  |
| Report an individual end point (not) included in the prior outcomes. | Yes ( ) No ( )  List if yes: specify the outcome, n, %) |
| Notes: | |

| **4. Available number of participants** | | |
| --- | --- | --- |
|  |  | Number |
| Baseline sample size | Yes ( ) No ( ) |  |
| Excluded from the baseline | Yes ( ) No ( ) |  |
| Lost to follow-up | Yes ( ) No ( ) |  |
| Other losses | Yes ( ) No ( ) |  |
| Total number included in the  analysis | Yes ( ) No ( ) |  |
| All subjects accounted for | Yes ( ) No ( ) |  |
| Notes: | | |

| **5. Statistical analysis** | |
| --- | --- |
| Statistical method used | Cox proportional hazards regression Yes ( ) No ( )  Other (specify): |
| Analysis includes males and females | Yes ( ) No ( ) |
| Study reports results by sex | Yes ( ) No ( ) |
| List all models used including unadjusted: |  |

_______

*Adapted from: Al Saikhan L, Park C, Hardy R, Hughes A. Prognostic implications of left ventricular strain by speckle-tracking echocardiography in population-based studies: a systematic review protocol of the published literature. BMJ Open. 2018 Jul 16;8(7):e023346. doi: 10.1136/bmjopen-2018-023346.*
